# Indicators of past COVID-19 infection status: Findings from a large occupational cohort of staff and postgraduate research students from a UK university

**DOI:** 10.1101/2020.12.07.20245183

**Authors:** Katrina A. S. Davis, Ewan Carr, Daniel Leightley, Valentina Vitiello, Gabriella Bergin-Cartwright, Grace Lavelle, Alice Wickersham, Michael H. Malim, Carolin Oetzmann, Catherine Polling, Sharon A.M Stevelink, Reza Razavi, Matthew Hotopf, On behalf of the KCL CHECK research team

**Author notes:** Corresponding author: Katrina A. S. Davis, Psychological Medicine, 3^rd^ Floor East, Institute of Psychiatry Psychology and Neuroscience Building, King’s College London (Denmark Hill Campus), London, SE5 8AZ, United Kingdom. joint last authors (Reza Razavi, Matthew Hotopf).

## Abstract

**Background:** Definitive diagnosis of COVID-19 requires resources frequently restricted to the severely ill. Cohort studies must rely on surrogate indicators to define cases of COVID-19 in the community. We describe the prevalence and overlap of potential indicators including self-reported symptoms, suspicion, and routine test results, plus home antibody testing.

**Methods:** An occupational cohort of 2807 staff and postgraduate students at a large London university. Repeated surveys covering March to June 2020. Antibody test results from ‘lateral flow’ IgG/IgM cassettes in June 2020.

**Results:** 1882 participants had valid antibody test results, and 124 (7%) were positive. Core symptoms of COVID-19 were common (770 participants positive, 41%), although fewer met criteria on a symptom algorithm (n=297, 16%). Suspicion of COVID-19 (n=509, 27%) was much higher than positive external tests (n=39, 2%). Positive antibody tests were rare in people who had no suspicion (n=4, 1%) or no core symptoms (n=10, 2%). In those who reported external antibody tests, 15% were positive on the study antibody test, compared with 24% on earlier external antibody tests.

**Discussion:** Our results demonstrate the agreement between different COVID indicators. Antibody testing using lateral flow devices at home can detect asymptomatic cases and provide greater certainty to self-report; but due to weak and waning antibody responses to mild infection, may under-ascertain. Multiple indicators used in combination can provide a more complete story than one used alone. Cohort studies need to consider how they deal with different, sometimes conflicting, indicators of COVID-19 illness to understand its long-term outcomes.

**THUMBNAIL:** *What is already known on this subject?:* Research into the effects of COVID-19 in the community is needed to respond to the pandemic, and guidance is needed as to how cohort studies measure COVID-19 infection status retrospectively, particularly given that objective testing for infection was not widely available in the first wave of COVID-19 in many countries. Retrospective testing might be possible using antibodies as a proxy for previous COVID-19 infection.

*What this study adds?:* Antibody testing is feasible in community cohorts but sensitivity may be poor. Self-report of suspected infection, recall of symptoms and results of tests received elsewhere add different aspects to the ascertainment of COVID-19 exposure. Combining self-report and objectively measured indicators may enable tailored algorithms for COVID-19 case definition that suits the aims of different research studies.

## INTRODUCTION

The majority of those affected by COVID-19 are community cases not requiring hospitalisation,[1] but research is needed about medium and long-term outcomes of community cases, particularly so-called “long COVID”.[2-6] Research in hospital cohorts has used a combination of clinical assessment, antigen testing by polymerase chain reaction (PCR), and lung imaging to provide a strong basis for COVID-19 diagnosis.[7] In community settings, such information is often unavailable. As these tests (particularly antigen/PCR tests) are time sensitive, participants in cohort studies may have missed the window where definitive diagnosis might be made. Since there are no “gold standard” methods by which community-based studies can distinguish between cases and controls, researchers rely on proxy indicators of COVID-19.

Potential indicators of past COVID-19 infection include (a) whether the participant thinks they have been infected; (b) report of symptoms of COVID-19; and (c) report of test results, all of which are dependent on participant recall. Detection of antibodies produced in response to SARS-CoV-2 (the virus that causes COVID-19) in blood samples may provide an objective measure of exposure, although it is not yet clear how long these antibodies remain detectable.[8] While there is guidance on clinical tests for COVID-19,[9, 10] there is little guidance for cohort studies for choosing and interpreting indicators in research. We provide a descriptive analysis of five potential indicators to appraise their use, using a cohort study of staff and postgraduate research students (PGRs) of a university in London, United Kingdom (UK), that has been running since April 2020.[11, 12]

## METHODS

Reporting conforms to The Strengthening the Reporting of Observational Studies in Epidemiology (STROBE) guidelines,[13] and a checklist can be found in appendix 1 of supplementary materials.

### Study

The King’s College London Coronavirus Health and Experiences of Colleagues at King’s (KCL-CHECK) study explores the health and wellbeing outcomes of the COVID-19 pandemic on staff and PGRs. Ethical approval has been gained from King’s Psychiatry, Nursing and Midwifery Research Ethics Committee (HR-19/20-18247). A protocol is available.[11] Briefly, eligible participants were current staff or PGRs residing in the UK (for antibody testing). All King’s staff and PGR students were invited to participate via email on April 16^th^ 2020 as well as advertisements on internal media / social media. The baseline survey was open for enrolment for two weeks. Participants provided informed consent and most opted into follow-up: 90% agreed to two-monthly surveys, 89% also agreed to shorter fortnightly surveys.

### Measures

Table 1 shows the schedule for follow-ups, with the first follow-up survey referred to as Period 1 (P1). Questions in the baseline and longer follow-up surveys asked about experiences in the last two months; questions in the shorter fortnightly surveys referred to the last two weeks. We report potential indicators from surveys at P0 (baseline) to P5 which took place between April and June 2020 and antibody testing in June 2020.

**Table 1.**
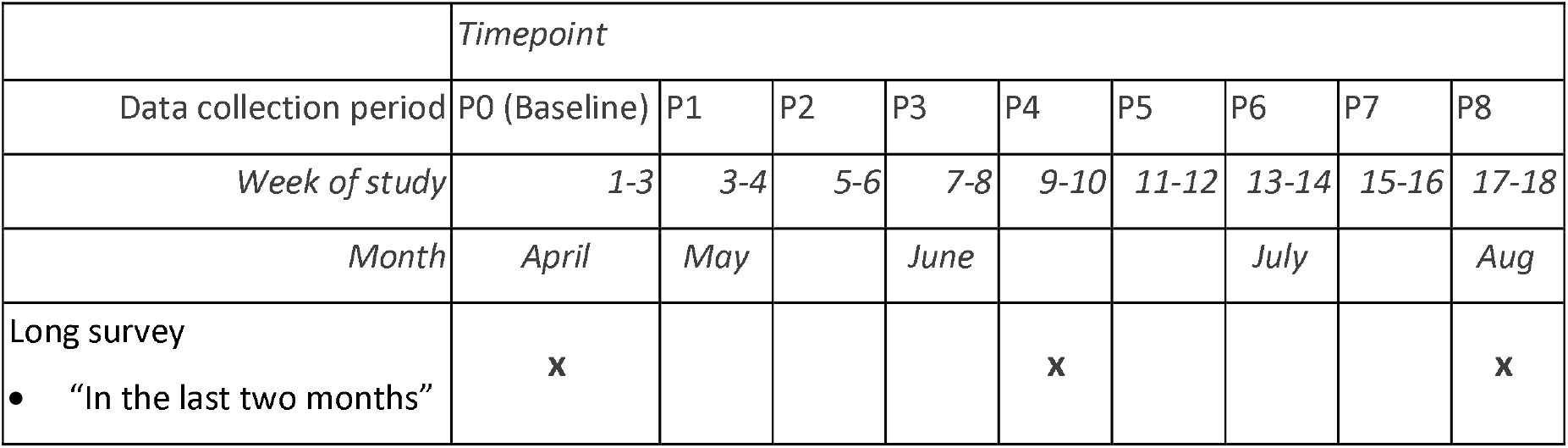
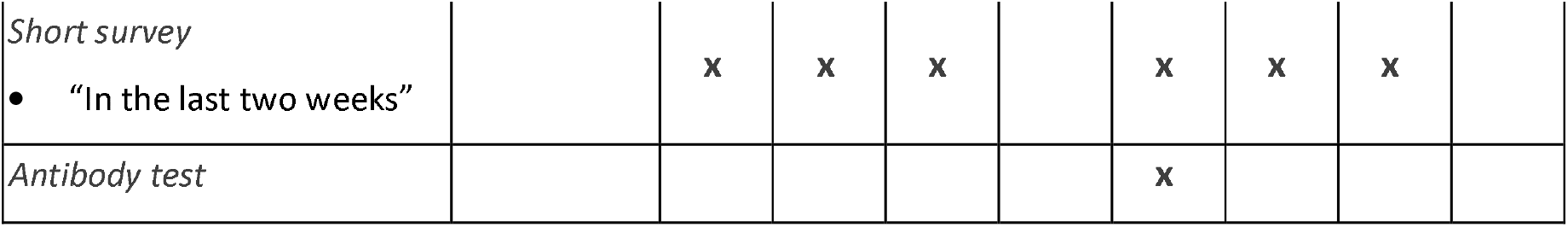
Periods of data collection for KCL CHECK to week 18 (April – Aug 2020).

|  | Timepoint |  |  |  |  |  |  |  |  |
| --- | --- | --- | --- | --- | --- | --- | --- | --- | --- |
| Data collection period | P0 (Baseline) | P1 | P2 | P3 | P4 | P5 | P6 | P7 | P8 |
| Week of study | 1-3 | 3-4 | 5-6 | 7-8 | 9-10 | 11-12 | 13-14 | 15-16 | 17-18 |
| Month | April | May |  | June |  |  | July |  | Aug |
| Long survey |  |  |  |  |  |  |  |  |  |
| • “In the last two months” | x |  |  |  | x |  |  |  | x |
| *Short survey* |  |  |  |  |  |  |  |  |  |
| • “In the last two weeks” |  | x | x | x |  | x | x | x |  |
| *Antibody test* |  |  |  |  |  | x |  |  |  |

#### Self-reported suspicion of COVID-19 illness

At the baseline (P0) participants were asked “Do you think that you have had COVID-19 (coronavirus) at any time? Definitely/Probably/Unsure/No”. At P1, P2, P3, and P5 participants were asked “Do you think that you have had COVID-19 (coronavirus) in the last two weeks?” At P4, participants were asked “Do you think that you have had COVID-19 (coronavirus) in the last two months?”. These responses were summarised as the highest degree of suspicion ever reported for each participant (across P0 to P5). For some analyses ‘Definitely’ and ‘Probably’ were combined to indicate positive suspicion of COVID-19.

#### Self-report of COVID-19 symptoms

We adapted the symptom list used by the ZOE coronavirus daily reporting app (part of the COVID symptom study) [14, 15] to cover two-month periods (P0 and P4) or two-week periods (P1, P2, P3 and P5), with a screening question: “In the last two months[weeks], how have you felt physically?”. We scored symptoms according to two definitions for comparison: (a) report of any ‘core symptom’ of fever, new persistent cough or loss of smell/taste; (b) the output of a symptom algorithm described by the COVID symptom study that incorporated loss of smell/taste, cough, severe fatigue and skipped meals, participant age and gender [15]. Symptoms were summarised as a binary indicator of whether participants, over all available survey periods, had ever vs. never (i) scored positively on the ZOE algorithm, (ii) reported a core symptom, (iii) reported feeling not right physically (‘any symptom’). If participants missed a survey period, they were considered to have not reported symptoms at that period.

#### Self-report of COVID-19 testing

We asked, “Have you had a test for COVID-19 (coronavirus)?” and “What was the result?” at baseline, and the same for two weeks at P1, P2, P3, and P5, and two months at P4. At P8, we asked again about past testing, distinguishing between a swab of the throat and/or nose to look for infection and a blood/blood spot test to look for evidence of past infection. These were used to split reported tests in P0 to P5 into “antigen/PCR tests” and “external antibody tests” (those without allocation at P8 were included as presumed antigen/PCR).

#### Home antibody tests

The SureScreen Diagnostics Rapid COVID-19 IgG/IgM Immunoassay Test Cassette was used to measure evidence of antibodies to the ‘spike’ protein of SARS-CoV-2. The performance of this test in laboratory conditions has been shown to be good, for example using samples from 268 keyworkers who self-reported positive COVID-19 antigen/PCR tests and 1,995 historical samples it had 94.0% sensitivity and 97.0% specificity, also showing 96.3% agreement with the SAR-CoV-2 spike antibody enzyme-linked immunoassay (ELISA) result for 2,847 keyworkers.[16] An internal pilot demonstrated that the test cassette could be used by participants without specific training, and following feedback received we developed our procedure and detailed illustrated instructions shown in supplementary material appendix 2. The test kit was posted to participants in late June, including the test cassette and a lancet for providing a blood spot. Participants were asked to upload a photograph of the result to a secure server. Participants could contact the team via email if they had difficulties, who answered within two working days, and could arrange for a replacement kit to be sent (sent in early July). The participant could rate the result as positive (IgG or IgM), negative or invalid, but this analysis uses only the rating given by the KCL-CHECK team who interpreted the photographs, as explained in a previous paper.[12]

#### Participant Characteristics

All characteristics were taken from the baseline survey. Ethnicity was asked using recommended wording from the Office of National Statistics with 18 groups,[17] but ethnicity is reported grouped into five categories due to small numbers of some ethnic groups. Role within the university was grouped into: (i) academic, specialist and management; (ii) research, clerical and technical; (iii) teaching, facilities and clinical; and (iv) PGR student. Participants were also asked if they were in roles that the government had designated as essential, making them a “keyworker”.

### Analysis

Datasets from each period and antibody testing were merged using R 4.0.0 and associated packages [18-21]. We summarised participation and missing data, then explored the overlap of indicators through descriptive analyses and figures. We give proportions to the nearest percentage point, unless under 1%, with 95% confidence intervals calculated using Wilson’s method. We calculated sensitivity and specificity of the self-report indicators compared to the KCL-CHECK antibody test, but do not necessarily regard the latter as a gold-standard for defining past COVID-19 cases.

## RESULTS

### Cohort and missing data

The baseline study included 2807 staff and PGRs, representing a response rate of 23% (Figure 1). The main analysis includes the 1882 participants who uploaded a valid antibody test by 13^th^ July (Figure 1, Table ST1). This cohort was 88% White, 71% female and 13% keyworkers, with a median age of 37 years. Tables ST2 and ST3 compare this cohort to known population characteristics of KCL staff and PGRs taken from King’s administrative data.

**Figure 1.**
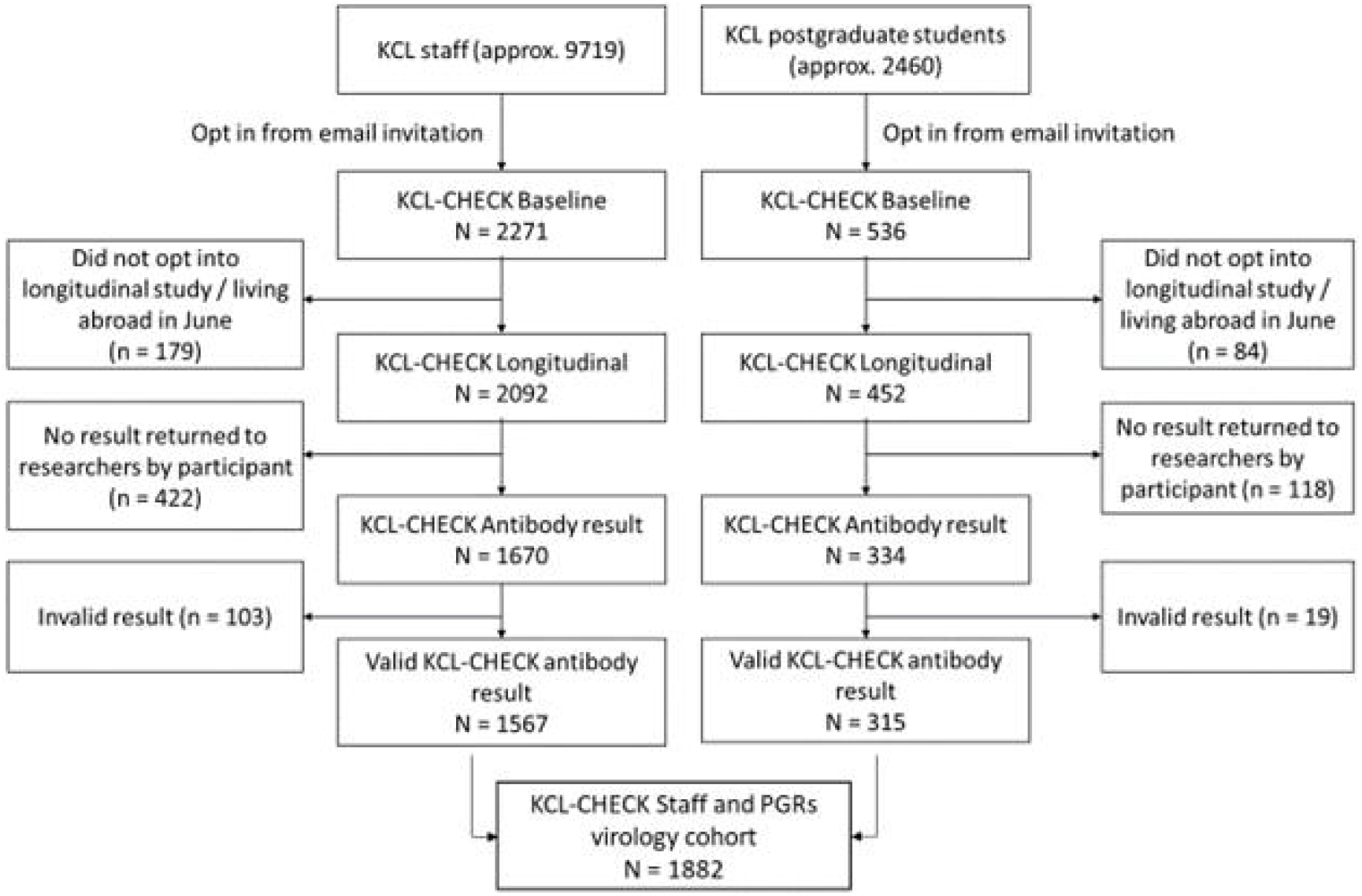
Study flowchart

Table 1 shows five opportunities to complete follow-up surveys before the antibody test: 98% completed at least one survey, 68% completed all five. As well as the analysis on 1882 participants, a secondary analysis limited to 1687 participants (90%) who had completing the P4 survey that asked about the two months from baseline to early June, thereby giving a near-complete overview of self-report prior to the antibody testing. Prevalence and overlap of COVID indicators were identical; the larger cohort is reported herein.

### COVID indicators

Table 2 shows the prevalence of COVID indicators in our sample. Of 1882, 124 (7%, 95% confidence interval 6-8) tested positive for antibodies. This compares with 814 (41%, CI 39-43) with at least one core symptom, 527 (27%, CI 25-29) suspecting they had experienced COVID-19 and 312 (16%, CI 14-18) positive on the symptom algorithm. Most who reported COVID indicators reported them at baseline, covering March-April 2020 (90% with core symptoms, 91% suspected, 88% symptom algorithm). 323 (17%, CI 16-19) reported receiving an external COVID-19 test: 235 antigen/PCR, 138 antibody, including 50 who had both. Thirty-nine reported a positive external test result (10/138 antigen, 33/235 antibody, including 4 both), 2% (CI 2-3) of all participants.

**Table 2.** The prevalence and overlap of positive COVID-19 indicators in KCL CHECK in order of prevalence in the main cohort (n=1882) with shading reflecting the strength of concordance.

|  | One or more core COVID-19 symptoms reported | Participant thinks they have had COVID-19 | Symptom algorithm positive | KCL CHECK antibody test positive | Reports positive test result from elsewhere |
| --- | --- | --- | --- | --- | --- |
| Overall prevalence | 770/1882, 41% | 509/1882, 27% | 297/1882, 16% | 124/1882, 7% | 39/1882, 2% |
| Number and proportion of column who also have: |  |  |  |  |  |
| One or more core COVID symptoms reported |  | 429 / 509 , 84% | 297 / 297 , 100% | 106 / 124 , 85% | 31 / 39 , 79% |
| Participant thinks they have had COVID | 429 / 770 , 56% |  | 214 / 297 , 72% | 101 / 124 , 81% | 33 / 39 , 85% |
| Symptom algorithm positive | 297 / 770 , 39% | 214 / 509 , 42% |  | 83 / 124 , 67% | 25 / 39 , 64% |
| KCL-CHECK antibody test positive | 106 / 770 , 14% | 101 / 509 , 20% | 83 / 297 , 28% |  | 24 / 39 , 62% |
| Reports positive test result from elsewhere | 31 / 770 , 4% | 33 / 509 , 6% | 25 / 297 , 8% | 24 / 124 , 19% |  |

The overlap between indicator results (Table 2) meant that meeting any one indicator increased the likelihood of all others. Of those with a core symptom of COVID-19, 56% thought they had experienced COVID-19, but fewer were positive for antibodies (14%) or an external test (4%). Of those who had tested positive on the KCL-CHECK antibody tests, 85% had experienced core symptoms, 81% thought they had had COVID-19 and 67% met the symptom algorithm. In the KCL-CHECK antibody positive group, 19% reported a positive external test result: a further 9% reported external tests but no positive test results. Concordance for positive and negative status is explored in Table ST4, ranging from 60% for core symptoms and external test, and 94% for KCL-CHECK antibody test and external test.

Table ST5 compares the external antibody test results and KCL-CHECK antibody test results for the 138 participants with external antibody test results. Agreement was 88% (kappa=0.64) with 24% (CI 18-32) of the participants positive on external antibody tests and 15% (CI 10-22) positive on KCL-CHECK antibody test.

Figure 2A and Table ST6 show that participants who thought they had not experienced COVID-19 were very unlikely to get a positive antibody test result (0.7%) with the proportion testing positive increasing to 39% among those who were definite. Probable or definite suspicion of COVID-19 infection had 81% sensitivity (101/124) and 77% specificity (1350/1758) for KCL-CHECK antibody test. Figure 2B shows that the majority of people who tested positive on the KCL-CHECK antibody test were positive on the symptom algorithm, which had 67% sensitivity (83/124) and 88% specificity (1544/1758) for KCL-CHECK antibody test. Core symptoms (including those also algorithm positive) had 85% sensitivity (106/124) and 62% specificity (1094/1758). Algorithm positive participants were six times as likely to test positively as those who only had core symptoms (Table ST6), but those with non-core symptoms were no more likely to test positive than those who reported no symptoms (2% non-core symptoms: 2% no symptoms).

**Figure 2.**
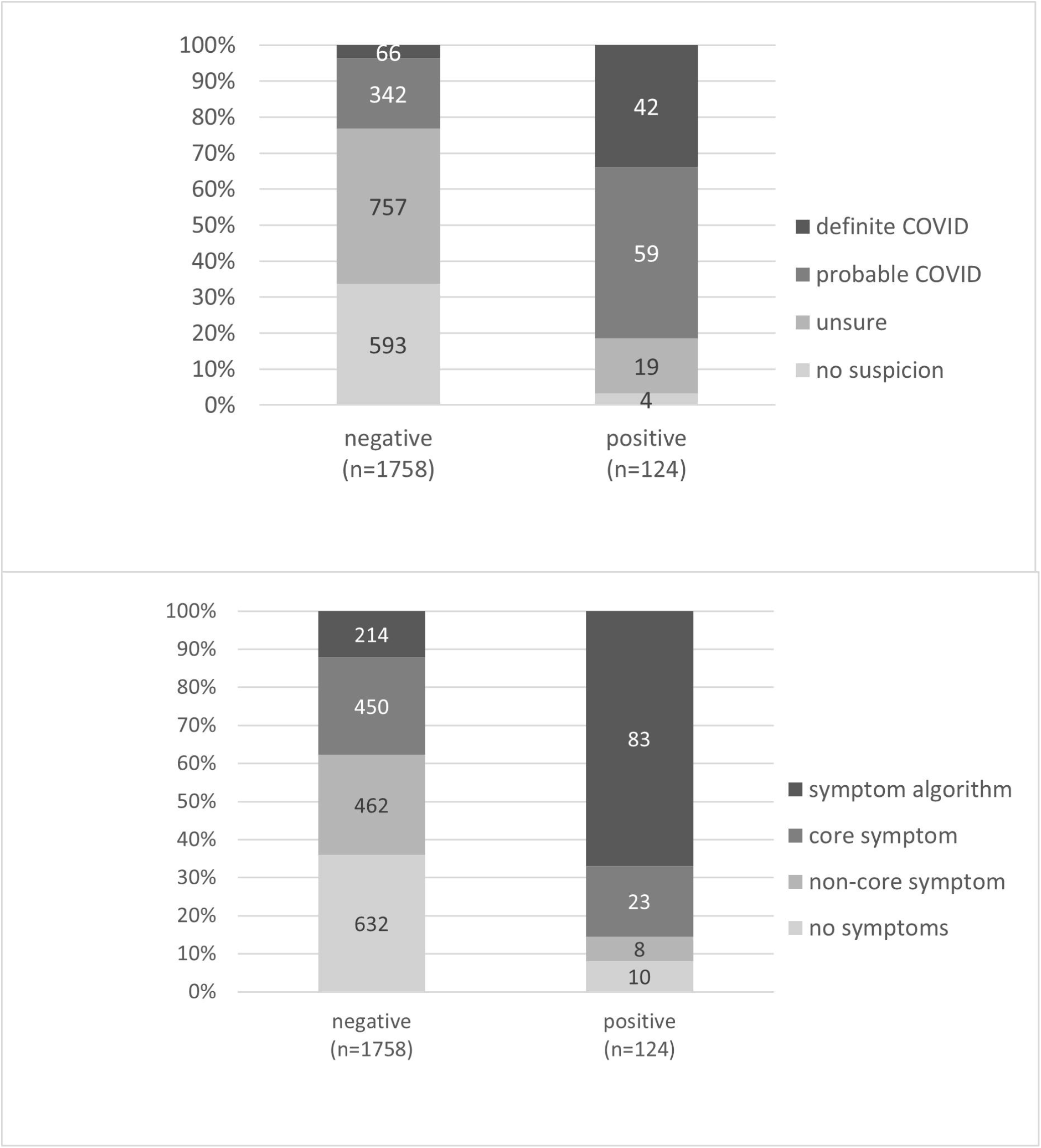
(A-B) KCL-CHECK antibody test result in June showing distribution of A. participant suspicion that they had experienced COVID-19 and B. highest level of symptoms reported, both between March and June2020. Participant numbers as labels

Table 3 shows participants at the intersect of each level of suspicion and symptom (algorithm, core, non-core, none). Where there were at least ten participants, the proportion testing positive on KCL-CHECK antibody test is given. For each level of symptoms, greater suspicion increased the probability of a positive KCL-CHECK antibody test, rising to 49% in those with algorithm symptoms and definite suspicion.

**Table 3.**
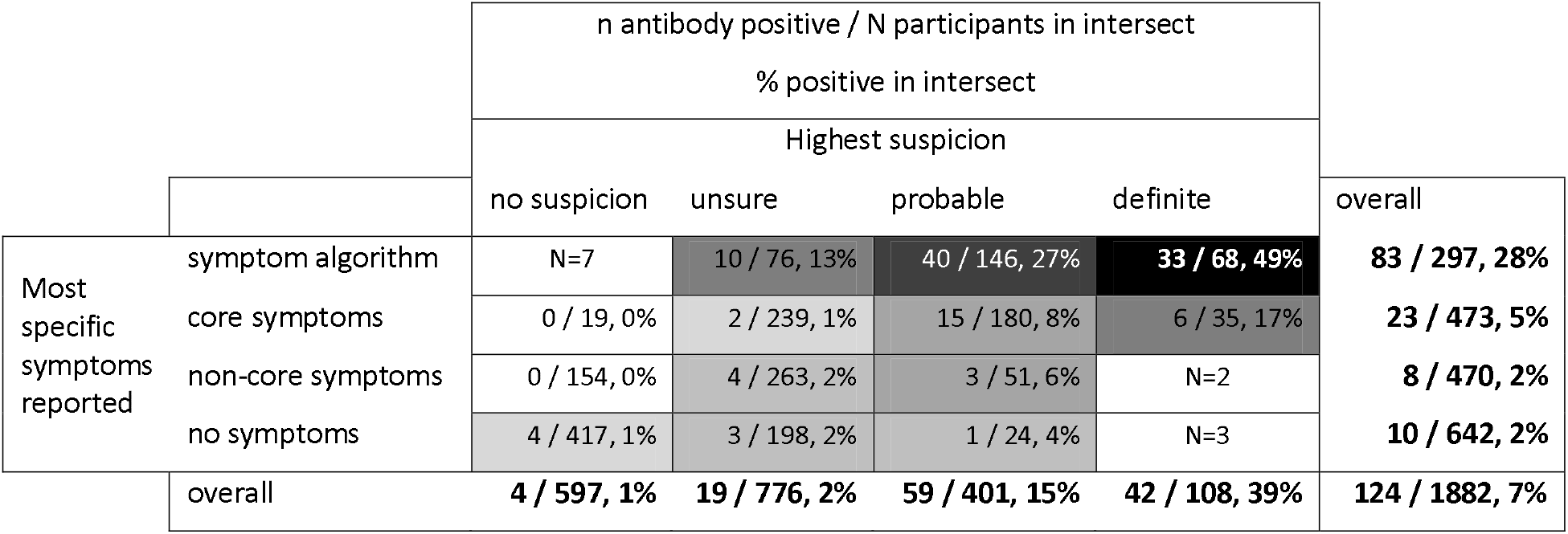
Proportion testing positive in intersecting groups of participant suspicion and self-reported symptoms (where at least 10 participants in the group). Depth of shading indicates proportion positive.

## DISCUSSION

Ascertaining cases and controls of COVID-19 in the community is challenging, given symptoms that are variable, common and non-specific, and testing in the first wave was not widely available.[1] However, community follow-up studies on the impact of COVID-19 illness are needed.[5, 6, 22] There are no “gold-standard” diagnostic criteria,[7] and for people who have not had the time-sensitive tests, there may be no opportunity to get diagnostic certainty. KCL CHECK used retrospective ascertainment based on self-report and home antibody testing to reduce the uncertainty – while acknowledging it could not recover the “ground truth”.

The KCL-CHECK cohort had, by June 2020, a prevalence of COVID indicators from 2% to 41%, depending on which one used. The 2% refers to self-reported positive external tests – but given that antigen testing was unavailable during the March 2020 wave of infections, and performance depends on timing and swab technique, this will under-estimate COVID-19.[23] The 41% refers to reporting one or more core COVID-19 symptoms, which overlap with other illnesses, over-estimating symptomatic COVID-19. While false positive suspicion of COVID-19 is likely to be high due to the high profile of the illness, we found it a useful indicator because low suspicion of COVID-19 was highly predictive of negative antibody testing, and higher suspicion added to the likelihood of testing positive in participants with similar symptoms. Presumably a participant’s suspicion factors in context such as symptom unusualness and contacts with COVID-19.

The COVID symptom study report that the algorithm we implemented was 65% sensitive and 78% specific for antigen/PCR test outcome on a single instance of scoring positive.[15] In a separate cohort (TwinsUK), ever being algorithm positive in March and April was 37% sensitive and 95% specific for laboratory antibody testing.[24] In our study, algorithm positivity in any of up to six surveys was 67% sensitive and 88% specific for home antibody test outcome. The high sensitivity of our symptom algorithm outcome shows that most people with antibodies had significant symptoms. In fact, our study found fewer asymptomatic people positive for antibodies (9% no symptoms, 15% no core symptoms) than many other studies, including TwinsUK (19% no symptoms, 27% no core symptoms)[24] and REACT2 (32% no symptoms, 39% no core symptoms).[25]

Antibody testing in KCL-CHECK used an IgG/IgM test kit based on “lateral flow” technology, sent to participants, which was simple to use and has high validity (under lab conditions).[26] Antibody tests can be fallible, giving false positives through cross-reactivity with antibodies unrelated to SARS-CoV-2, giving false positive results approximately 2 per 100,[10, 16] which can be problematic in large studies with low prevalence. Sensitivity is also a concern, since small numbers of people do not produce anti-spike antibodies,[27, 28] and they are detected more inconsistently in mild COVID-19,[26, 29, 30] and may decline over time.[31-33]. Testing in KCL-CHECK occurred at least three months after most participants’ symptoms: few studies have tracked antibody levels over this time. Antibodies may cease to be detectible months after exposure, especially on lateral flow devices.[34, 35] Among KCL-CHECK participants who reported previous antibody testing, 15% were positive on the KCL-CHECK antibody test, compared with 24% in their prior reported test. This may suggest time-dependent loss of reactivity, although there were likely also differences in test specifications and small numbers. Further rounds of testing of our cohort may help clarify this. Augmenting antibody testing with testing for T cell response to better track long-term immunity may be possible in the future.[36]

For other cohort studies, our results suggest that collecting results of external testing will underestimate the proportion infected. Collecting symptom report for the COVID symptom study algorithm [15] will assist in finding those who have had a COVID-19-like illness, and considering the participants’ own suspicion can add both sensitivity and specificity. Testing with a high specificity antibody test will identify past cases that were asymptomatic or atypically symptomatic and add more certainty where a positive test accords with COVID-19 symptoms. However, timing may lead to poor sensitivity when testing is used in isolation.

### Strengths and weaknesses

The strengths of this study include the survey repeating every fortnight to minimise recall bias. We incorporated a symptom checklist that has been previously evaluated. The antibody test kit was highly specific for SARS-CoV-2, suited to minimise false positives in population screening. While our conclusions could have been strengthened by the presence of a hospital standard diagnosis against which to compare other outcomes, the paper aimed to show what results can be gathered in the community.

Home testing maximised uptake of the test at a time when people may have been hesitant about attending a clinic. The lateral flow cassette is designed for use by a trained person but, from our pilot and the high proportion of people returning valid results, we believe that with illustrated instructions and a responsive email enquiry address most participants were able to perform the test.[12] Nevertheless, the potential for errors and inconsistencies when carrying out tests out of the laboratory.[16, 30]

The analysis utilised results from all with valid antibody testing, regardless of survey completion. Our sensitivity analysis showed that restricting to a more complete sample made little difference, possibly because COVID-19 infections were much less common in May/June 2020 than they had been in March,[37, 38] so we would expect relatively few positives to occur after the April baseline.

Finally, our cohort comprised staff and PGRs from a single university, with over-representation of women, people of White ethnicity and those in management/research roles. This was not representative of the general population. We expect that our experience will be useful to studies that are of different composition in the community, although care would need to be taken in populations with very different expected COVID-19 prevalence.

## Conclusions and implications

This paper shows a variety of potential COVID-19 indicators that may be available for community studies, their prevalence and overlap in a single cohort in the challenging context of COVID-19 research: the time-course of detectable antigen and antibody, poor access to routine testing at times, symptoms that overlap with many other illnesses, and the high profile of the illness. We show that participant suspicion is a useful adjunct to symptom report, although false positives can be expected. Adding the symptom algorithm may increase specificity, and an antibody test may add asymptomatic cases that would otherwise be missed. We encourage researchers to consider the use of algorithms that maximise COVID-19 case history, rather than relying on single measures which may give a false sense of certainty.

## Data Availability

Researchers may access pseudonymised data by application to the Principal Investigators (Professor Matthew Hotopf, Professor Reza Razavi and Dr Sharon Stevelink) subject to conditions set out in the protocol

## Acknowledgements

The research team would like to thank the participants of KCL-CHECK and the team that has provided logistic and advisory support, in particular: Jonathan Edgeworth, Liam Jones, Lisa Sanderson, Jana Kim, Laila Danesh, Charlotte Williamson, Laurence Blight, Rupa Bhundia, Lucy O’Neill, Candice Middleton. We also thank Mark Zuckerman for comments on an earlier draft of this paper. This paper represents independent research supported by the National Institute for Health Research (NIHR) Biomedical Research Centre at South London and Maudsley NHS Foundation Trust and King’s College London. The views expressed are those of the author(s) and not necessarily those of the NHS, the NIHR or the Department of Health and Social Care. MHM is a Wellcome Trust Investigator.

## Data availability

Researchers may access pseudonymised data by application to the Principal Investigators (Professor Matthew Hotopf, Professor Reza Razavi and Dr Sharon Stevelink) subject to conditions set out in the protocol https://www.medrxiv.org/content/10.1101/2020.06.16.20132456v2.

## Supplemental material

Supplementary Table ST1 Cohort characteristics at different points in the study, with the “valid antibody result” cohort being used in this paper

Supplementary Table ST2 Characteristics PGR students

Supplementary Table ST3 Cohort characteristics staff

Supplementary Table ST4 Overall agreement of COVID status between pairs of COVID-19 outcomes (positive and negative) as a proportion

Supplementary Table ST5 Comparing external antibody testing results with KCL-CHECK antibody test result (in those with external result)

Supplementary Table ST6 Comparing participant reported suspicion of COVID-19 illness (highest suspicion reported April-June) with KCL-CHECK antibody testing result in June

Supplementary Table ST7 Comparing participant reported symptoms (most specific reported April-June) with KCL-CHECK antibody testing result in June

Appendix 1: STROBE checklist

Appendix 2: Home testing procedure and instructions

