## Supplemental Material for "Indicators of past COVID-19 infection status: Findings from a large occupational cohort of staff and postgraduate research students from a UK university"

Supplementary Tables (ST1 – ST7)

Appendix 1: STROBE checklist

Appendix 2: Home testing procedure and instructions

Supplementary Table ST1 Cohort characteristics at different points in the study, with the "valid antibody result" cohort being used in this paper
Supplementary Table ST2 Characteristics PGR students
Supplementary Table ST3 Cohort characteristics staff
Supplementary Table ST4 Overall agreement of COVID status between pairs of COVID-19 outcomes (positive and negative) as a proportion
Supplementary Table ST5 Comparing external antibody testing results with KCL-CHECK antibody test result (in those with external result)
Supplementary Table ST6 Comparing participant reported suspicion of COVID-19 illness (highest suspicion reported April-June) with KCL-CHECK antibody testing result in June
Supplementary Table ST7 Comparing participant reported symptoms (most specific reported April-June) with KCL-CHECK antibody testing result in June

*Supplementary Table ST1 Cohort characteristics at different points in the study, with the "valid antibody result" cohort being used in this paper*

| Cohort name (see figure 1) | | Baseline | Longitudinal | Antibody result | Valid antibody result |
|---|---|---|---|---|---|
| Gender | Female | 1948 (69%) | 1769 (70%) | 1418 (71%) | 1339 (71%) |
| | Male | 842 (30%) | 760 (30%) | 574 (29%) | 533 (28%) |
| | Other | 17 (1%) | 15 (1%) | <10 (1%)* | <10 (1%)* |
| Role | Academic, specialist and management. | 1075 (38%) | 1060 (42%) | 866 (43%) | 810 (43%) |
| | Research, clerical and technical | 809 (29%) | 797 (31%) | 661 (33%) | 622 (33%) |
| | Teaching, facilities and clinical | 204 (7%) | 199 (8%) | 142 (7%) | 134 (7%) |
| | Post Graduate Research Students | 536 (19%) | 452 (18%) | 334 (17%) | 315 (17%) |
| | missing | 183 (7%) | 36 (1%) | <10 (1%)* | <10 (1%)* |
| Age-group (years) | 18-24 | 136 (5%) | 125 (5%) | 93 (5%) | 90 (5%) |
| | 25-34 | 1084 (39%) | 948 (37%) | 732 (37%) | 695 (37%) |
| | 35-44 | 762 (27%) | 698 (27%) | 566 (28%) | 535 (28%) |
| | 45-54 | 440 (16%) | 409 (16%) | 316 (16%) | 287 (15%) |
| | 55-64 | 307 (11%) | 288 (11%) | 239 (12%) | 220 (12%) |
| | 65+ | 78 (3%) | 76 (3%) | 58 (3%) | 55 (3%) |
| Ethnicity group | White | 2152 (77%) | 695 (27%) | 535 (27%) | 287 (15%) |
| | Mixed | 102 (4%) | 46 (1.8%) | 34 (1.7%) | 29 (1.5%) |
| | Asian | 204 (7%) | 191 (7.5%) | 133 (6.6%) | 125 (6.6%) |
| | Black | 39 (1%) | 37 (1.5%) | 21 (1%) | 20 (1.1%) |
| | Other | 70 (2%) | 69 (2.7%) | 49 (2.4%) | 44 (2.3%) |
| | missing | 240 (9%) | 23 (1%) | <10 (1%)* | <10 (1%)* |
| Keyworker status | Keyworker | 358 (13%) | 357 (14%) | 257 (13%) | 242 (13%) |
| | Non-keyworker | 2449 (87%) | 2187 (86%) | 1747 (87%) | 1640 (87%) |
| Total | | 2807 | 2544 | 2004 | 1882 |

*True value supressed due to small cell size

*Supplementary Table ST2 Characteristics PGR students*

| | | Approximate sampling frame (from KCL Student Office) | Post-Graduate student cohort |
|---|---|---|---|
| Gender | Female | 1390 (57%) | 229 (73%) |
| | Male | 1070 (43%) | 83 (26%) |
| Age-group | 18-24 | 690 (28%) | 50 (16%) |
| | 25-34 | 1315 (53%) | 199 (63%) |
| | 35+ | 455 (18%) | 66 (21%) |
| Ethnicity group | White | 1454 (59%) | 248 (79%) |
| | Mixed | 130 (5%) | <10 (2%*) |
| | Asian | 556 (23%) | 37 (12%) |
| | Black | 84 (3%) | <10 (2%*) |
| | Other | 170 (7%) | 16 (5%) |
| Total | | 2460 | 315 |

* True value suppressed due to small cell size

*Supplementary Table ST3 Cohort characteristics staff*

| | | Approximate sampling frame (from KCL Human Resources) | Staff cohort |
|---|---|---|---|
| Gender | Female | 5351 (55%) | 1110 (71%) |
| | Male | 4368 (45%) | 450 (29%) |
| Age-group | 18-24 | 464 (5%) | 40 (3%) |
| | 25-34 | 3374 (35%) | 496 (32%) |
| | 35-44 | 2755 (28%) | 495 (32%) |
| | 45-54 | 1794 (18%) | 276 (18%) |
| | 55-64 | 1084 (11%) | 207 (13%) |
| | 65+ | 248 (3%) | 53 (3%) |
| Ethnicity group | White | 6779 (70%) | 1412 (90%) |
| | Mixed | 415 (4%) | 23 (1%) |
| | Asian | 1212 (12%) | 88 (6%) |
| | Black | 520 (5%) | 14 (1%) |
| | Other | 793 (8%) | 28 (2%) |
| Total | | 9719 | 1567 |

*Supplementary Table ST4 Overall agreement of COVID-19 status between pairs of COVID indicators (positive and negative) as a proportion*

| Complete cohort (n=1882) | Core symptoms | Participant suspicion of COVID | Symptom algorithm | KCL -CHECK antibody test | External testing |
|---|---|---|---|---|---|
| Proportion with positive outcome | 41% | 27% | 16% | 7% | 2% |
| Agreement: | % (n agree) | % (n agree) | % (n agree) | % (n agree) | % (n agree) |
| Core | | | | | |
| Suspicion | 78% (1461) | | | | |
| Algorithm | 75% (1409) | 80% (1504) | | | |
| KCL -CHECK antibody test | 64% (1200) | 77% (1451) | 86% (1626) | | |
| External test | 60% (1135) | 74% (1400) | 85% (1596) | 94% (1767) | |

*Supplementary Table ST5 Comparing external antibody testing results with KCL-CHECK antibody test result (in those with external result)*

| | | External antibody testing | | | | |
|---|---|---|---|---|---|---|
| | | Negative | Positive | Total | | |
| KCL-CHECK antibody test result | Negative | 103 | 14 | 117 | 2x2 stats | |
| | Positive | 2 | 19 | 21 (15%) | agreement | 88% |
| | Total | 105 | 33 (24%) | 138 | (chance agreement) | 68%) |
| | | | | | kappa | 0.636 |
| Sensitivity, specificity, PPV and NPV refer to the ability of the KCL-CHECK antibody test to detect those who have tested positive on an external antibody test | | | | | sensitivity | 58% |
| | | | | | specificity | 98% |
| | | | | | Positive predictive value | 90% |
| | | | | | Negative predictive value | 88% |

*Supplementary Table ST6 Comparing participant reported suspicion of COVID-19 illness (highest suspicion reported April-June 2020) with KCL-CHECK antibody testing result in June 2020*

| Participant suspicion COVID-19 infection | KCL-CHECK antibody **negative** | KCL-CHECK antibody **positive** | %positive | Prevalence ratio, relative to no suspicion |
|---|---|---|---|---|
| No suspicion (n=597) | 593 | 4 | 0.7% | ref |
| Unsure (n=776) | 757 | 19 | 2.4% | 3.7 |
| Probable COVID (n=401) | 342 | 59 | 14.7% | 22.0 |
| Definite COVID (n=108) | 66 | 42 | 38.9% | 58.0 |
| Total (n=1882) | 1758 | 124 | 6.6% | -- |

*Supplementary Table ST7 Comparing participant reported symptoms (most specific reported April-June 2020) with KCL-CHECK antibody testing result in June 2020*

| | KCL-CHECK antibody **negative** | KCL-CHECK antibody **positive** | %positive | Prevalence ratio, relative to no symptoms |
|---|---:|---:|---:|---:|
| No symptom (n=642) | 632 | 10 | 1.6% | ref |
| Non-core symptom (n=470) | 462 | 8 | 1.7% | 1.1 |
| Core symptom (n=473) | 450 | 23 | 4.9% | 3.1 |
| Symptom algorithm positive (n=297) | 214 | 83 | 27.9% | 17.9 |
| Total (n=1882) | 1758 | 124 | 6.6% | -- |

### Appendix 1:

STROBE Statement—Checklist of items that should be included in reports of ***cohort studies***

| | Item No | Recommendation | Section |
|---|---|---|---|
| **Title and abstract** | 1 | (*a*) Indicate the study's design with a commonly used term in the title or the abstract | Title |
| | | (*b*) Provide in the abstract an informative and balanced summary of what was done and what was found | Abstract |
| **Introduction** | | | |
| Background/rationale | 2 | Explain the scientific background and rationale for the investigation being reported | 1 |
| Objectives | 3 | State specific objectives, including any prespecified hypotheses | 1 (descriptive) |
| **Methods** | | | |
| Study design | 4 | Present key elements of study design early in the paper | 2.1-2.2 |
| Setting | 5 | Describe the setting, locations, and relevant dates, including periods of recruitment, exposure, follow-up, and data collection | 2.1 (and separate protocol) |
| Participants | 6 | (*a*) Give the eligibility criteria, and the sources and methods of selection of participants. Describe methods of follow-up | 2.1 (and separate protocol) |
| | | (*b*) For matched studies, give matching criteria and number of exposed and unexposed | n/a |
| Variables | 7 | Clearly define all outcomes, exposures, predictors, potential confounders, and effect modifiers. Give diagnostic criteria, if applicable | 2.2 |
| Data sources/ measurement | 8* | For each variable of interest, give sources of data and details of methods of assessment (measurement). Describe comparability of assessment methods if there is more than one group | 2.2 |
| Bias | 9 | Describe any efforts to address potential sources of bias | n/a |
| Study size | 10 | Explain how the study size was arrived at | n/a |
| Quantitative variables | 11 | Explain how quantitative variables were handled in the analyses. If applicable, describe which groupings were chosen and why | 2.2-2.3 |
| Statistical methods | 12 | (*a*) Describe all statistical methods, including those used to control for confounding | 2.3 |
| | | (*b*) Describe any methods used to examine subgroups and interactions | n/a |
| | | (*c*) Explain how missing data were addressed | 2.3-3.1 |
| | | (*d*) If applicable, explain how loss to follow-up was addressed | 3.1 |
| | | (*e*) Describe any sensitivity analyses | 2.3-3.1 |
| **Results** | | | |
| Participants | 13* | (a) Report numbers of individuals at each stage of study—eg numbers potentially eligible, examined for eligibility, confirmed | Figure 1 / Table ST1 |

| | | | |
|---|---|---|---|
| | | eligible, included in the study, completing follow-up, and analysed | |
| | | (b) Give reasons for non-participation at each stage | Figure 1 |
| | | (c) Consider use of a flow diagram | Figure 1 |
| Descriptive data | 14* | (a) Give characteristics of study participants (eg demographic, clinical, social) and information on exposures and potential confounders | Tables ST1-3 |
| | | (b) Indicate number of participants with missing data for each variable of interest | Table ST1, section 3.1 |
| | | (c) Summarise follow-up time (eg, average and total amount) | Section 3.1 |
| Outcome data | 15* | Report numbers of outcome events or summary measures over time | Table 2 |
| Main results | 16 | (*a*) Give unadjusted estimates and, if applicable, confounder-adjusted estimates and their precision (eg, 95% confidence interval). Make clear which confounders were adjusted for and why they were included | 3.2 |
| | | (*b*) Report category boundaries when continuous variables were categorized | 2.2 |
| | | (*c*) If relevant, consider translating estimates of relative risk into absolute risk for a meaningful time period | n/a |
| Other analyses | 17 | Report other analyses done—eg analyses of subgroups and interactions, and sensitivity analyses | 3.1 |
| **Discussion** | | | |
| Key results | 18 | Summarise key results with reference to study objectives | 4 |
| Limitations | 19 | Discuss limitations of the study, taking into account sources of potential bias or imprecision. Discuss both direction and magnitude of any potential bias | 4 |
| Interpretation | 20 | Give a cautious overall interpretation of results considering objectives, limitations, multiplicity of analyses, results from similar studies, and other relevant evidence | 4 |
| Generalisability | 21 | Discuss the generalisability (external validity) of the study results | 4 |
| **Other information** | | | |
| Funding | 22 | Give the source of funding and the role of the funders for the present study and, if applicable, for the original study on which the present article is based | Funding[a] |

Table note (a): KCL-CHECK is funded by King's College London. All investigators are employed by King's College London. Two of the principal investigators are also managers in King's College London. Decisions regarding study design, analysis, interpretation and decision to publish have been taken by the named investigators without external influence.

*Give information separately for exposed and unexposed groups.

**Note:** An Explanation and Elaboration article discusses each checklist item and gives methodological background and published examples of transparent reporting. The STROBE checklist is best used in conjunction with this article (freely available on the Web sites of PLoS Medicine at http://www.plosmedicine.org/, Annals of Internal Medicine at http://www.annals.org/, and Epidemiology at http://www.epidem.com/). Information on the STROBE Initiative is available at http://www.strobe-statement.org.

#### Appendix 2: PDF of instructions sent to participants for home testing of antibodies

Dear «Follow_3»,

Thank you for your participation in the KCL CHECK (Coronavirus: Health & Experiences of Colleagues at King's) study. You are contributing to research concerning an unprecedented situation and are helping to improve understanding of how the COVID-19 (Coronavirus) pandemic is affecting the work, health and wellbeing of staff and postgraduate research students at King's College London.

As part of KCL CHECK, you consented to taking part in the virology component of the study and we are pleased to enclose your third antibody home-testing kit. We are repeating antibody testing a number of times throughout the study period, to assess antibody status over time and how this may change.

This test is comprised of a simple device used to detect the presence or absence of two types of COVID-19 antibodies (IgG and IgM) in a blood sample. Studies have shown the body produces these antibodies after experiencing COVID-19 infection. However, this is a research study and not a clinical test, so you should not rely only on the result of this test as either confirming that you have not had COVID-19 if the test is negative, or that you have had COVID-19 if the test is positive.

The test involves pricking your fingertip and taking a small blood sample which will be analysed in the test cassette supplied. This process should take around 10 minutes. You will need to photograph the result within 10-15 minutes of completing the procedure and upload the picture to the KCL CHECK website: www.kcl-check.org/test-results.

Enclosed you will find:

1. A Coronavirus Rapid Test Cassette and kit (contents explained further in the instructions);
2. Step-by-step instructions for how to perform the test, how to interpret the result, and how to share the result with the KCL CHECK Research Team. **Please read these before conducting the test and follow the steps carefully.**

Please note that we will be unable to provide replacement testing kits on this occasion. Therefore, we encourage you to please follow these instructions carefully before proceeding. You can find more information about the KCL CHECK study on the Participant Information Sheet at www.kcl-check.org/pis. The FAQ section of our website can also be found here: www.kcl-check.org/faq. If you have any further questions or if you would like to speak to the KCL CHECK Research Team, please.

Thank you for participating in this important research.

Best wishes,

**KCL CHECK Research Team**

**Please turn over for the step-by-step instructions when you are ready to begin the test**

Participant ID: «Login_ID»

**KCL CHECK**
**Instructions for COVID-19 antibody home-testing kit**
**(COVID-19 IgG/IgM Rapid Test Cassette)**

**Test kit contents:**

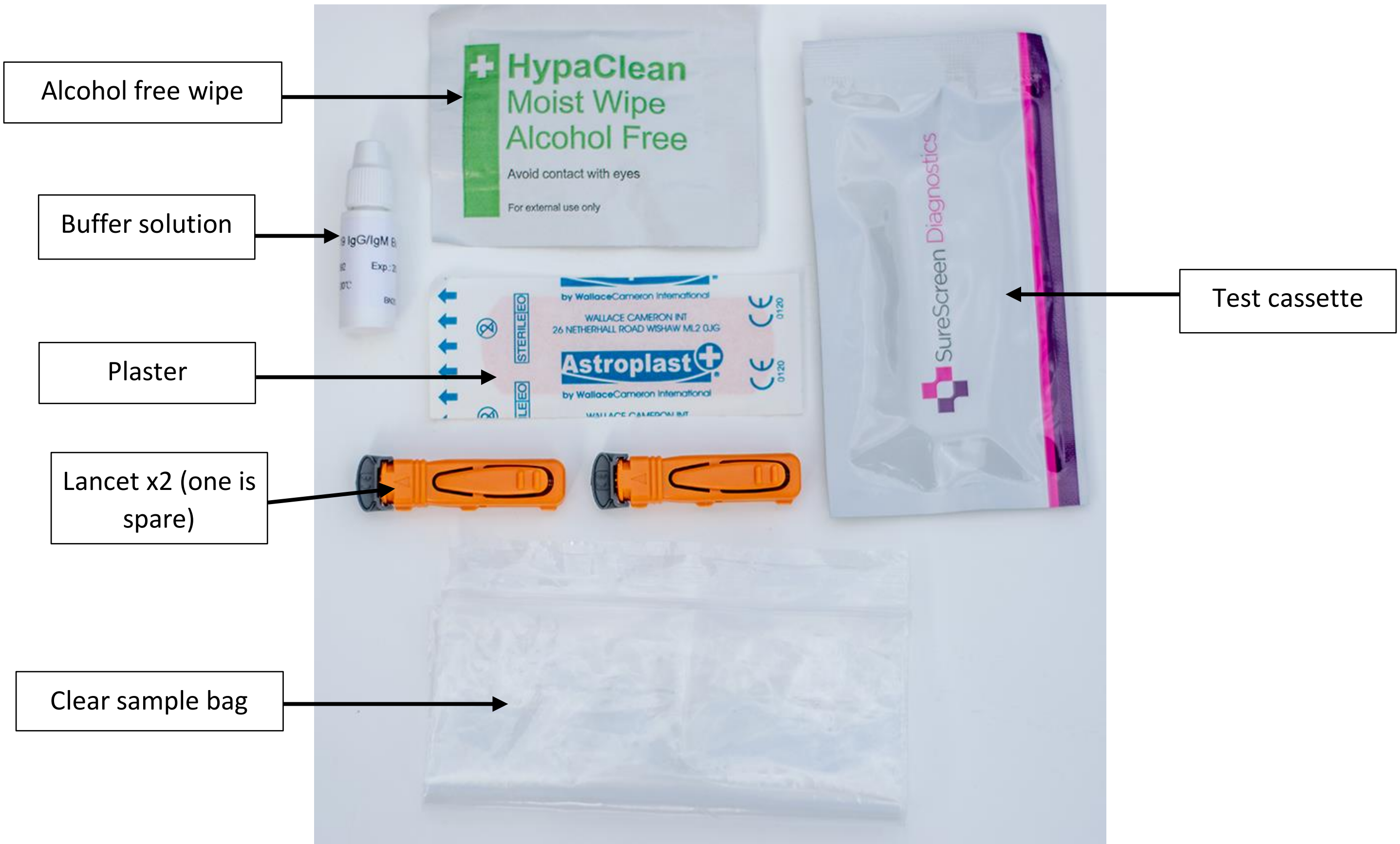


You also need a **<u>clean tissue</u>**, a **<u>timer</u>**, a **<u>pen</u>** and a way **<u>to photograph</u>** your result.

#### Step-by-step instructions

**The test must be interpreted and photographed within <u>10-15</u> minutes of completing the procedure to work correctly, so make sure you have time before starting.**

##### Step 1: Set-up

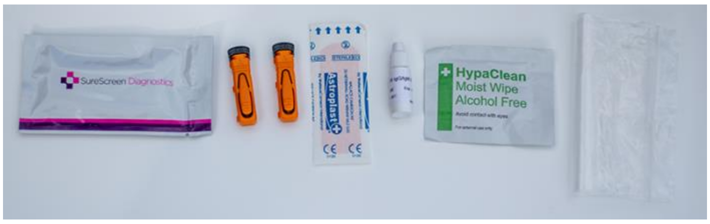


Before you begin, check you have everything you need (use the contents image above for reference). Then, remove the test cassette from the packaging **(use within 1 hour of opening)** and lay out the contents of your test kit on a clean, level surface. **Please note the buffer solution could be provided in a small, transparent vial instead of a screw-cap bottle.**

**Step 2: Wash your hands and wipe your fingertip**

Using warm water, wash your hands. This helps with blood flow. Then, select either your middle or ring finger, and use the alcohol-free wipe to clean it.

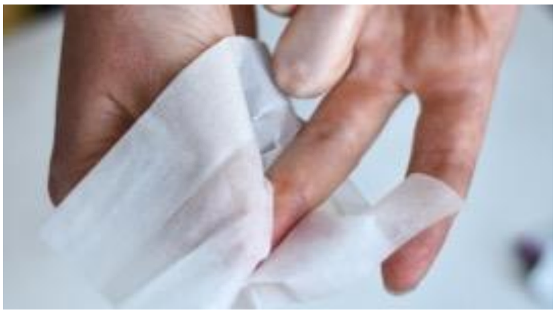

**Step 3: Puncture the skin using a lancet**

To use the lancet, hold the case from the sides and **twist** the cap a quarter turn to unlock. You will feel the cap advance forward; **do not pull**. Continue twisting until you feel it separate from the device then pull the cap. Use the second lancet if you have problems with the first.

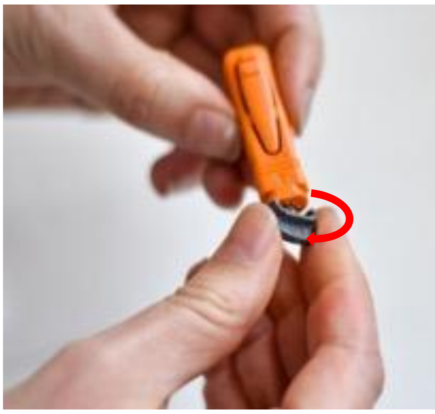

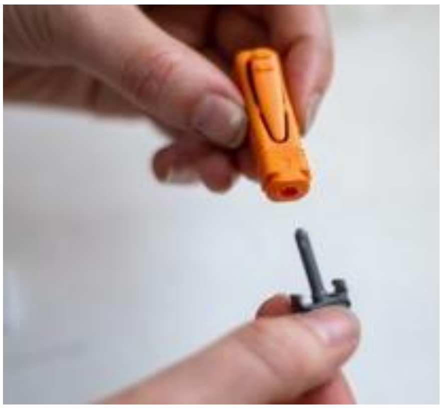

With the cap removed, massage your hand without touching the puncture site by rubbing down the hand towards the fingertip to encourage blood flow.

Then, hold the lancet firmly on your fingertip, and press the button to release the needle. The lancets are **single use** therefore please ensure that they are pressed firmly on the finger before firing.

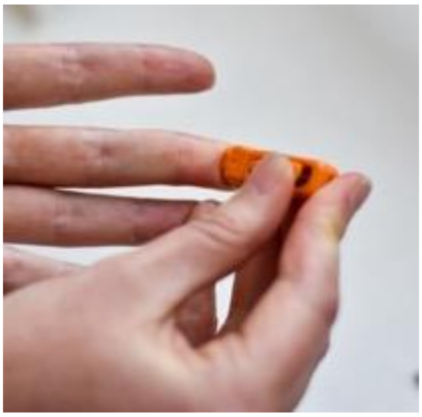

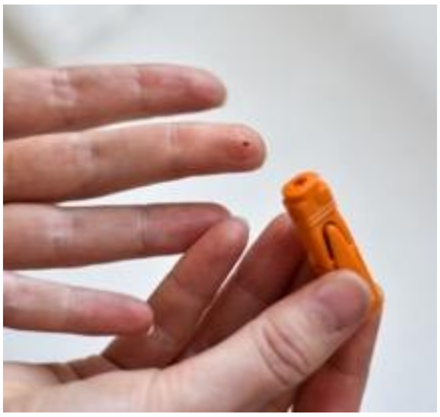

Wipe away the first sign of blood with a clean tissue.

**Step 4: Add a drop of blood to the test cassette**

Gently run your hand from the wrist up to the finger to form a rounded drop of blood. Release one drop of blood into the specimen well of the test cassette, **<u>this is marked 'S'</u>**.

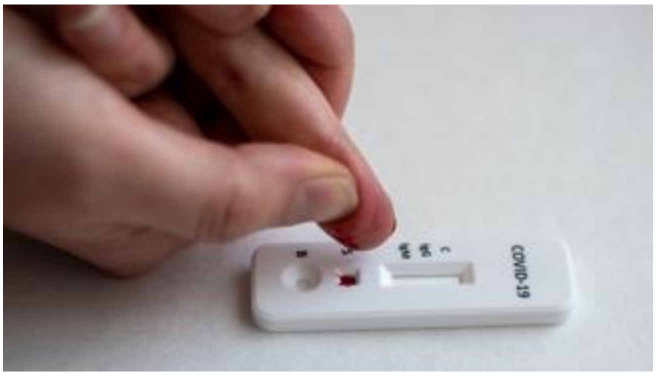


**Step 5: Add the buffer to the test cassette**

Open the buffer tube and add the buffer onto the test cassette area **<u>marked 'B'</u>** by squeezing. You must **<u>add two-three drops of buffer</u>**, try to do it as quickly as possible.

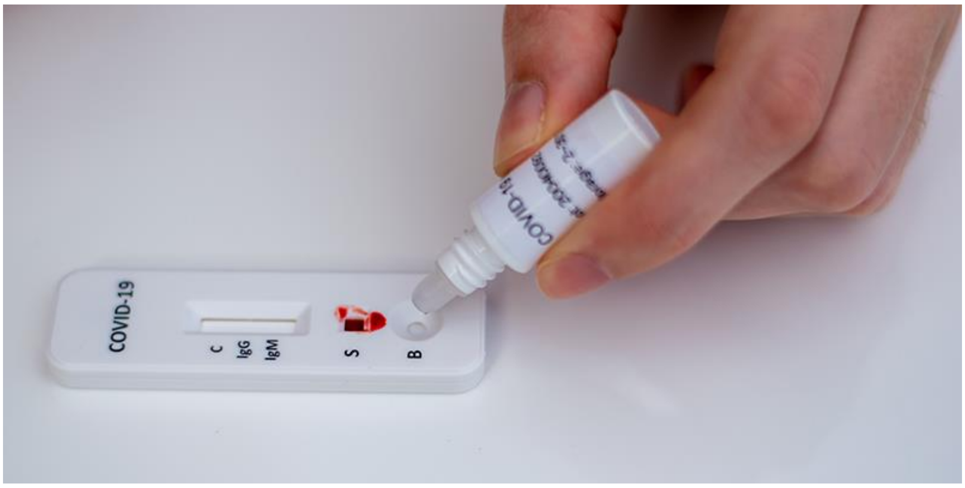

**Step 6: Start the timer and record the current time**

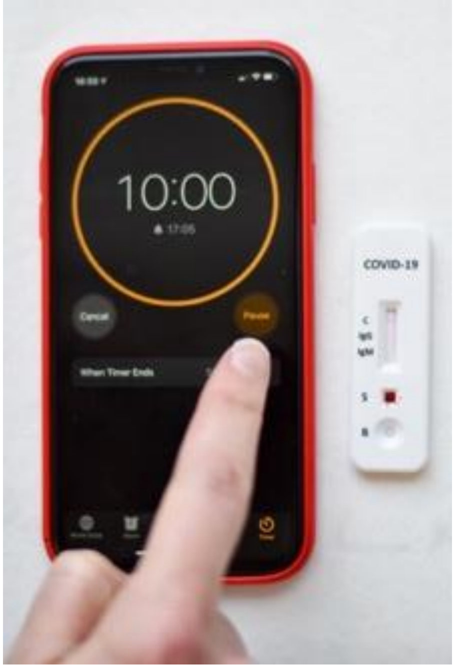


As soon as you have added the buffer, start a timer for **10 minutes**. Check the current time and record it in the box on the next page as the ‘test start time’. Please record the time in a 24-hour format (e.g.16:55).

While you wait, use the plaster to seal the puncture site on your fingertip.

**Step 7: Check your result**

After **10 minutes**, coloured line(s) should appear on the test cassette. You can use the guide below to check your results.

**Result Interpretation Guide:** (See Figure 1)

**Note: The COVID-19 IgG/IgM Rapid Test Cassette will only indicate the presence of COVID-19 antibodies and should not be used as the <u>sole</u> criteria for the diagnosis of COVID-19. A negative result does not at <u>any time</u> exclude the possibility of COVID-19 infection. Similarly, a positive result <u>does not</u> confirm a previous COVID-19 infection. Please follow current NHS guidelines regarding COVID-19 symptoms and diagnosis.**

**IgG and IgM POSITIVE:** Three lines appear. A colored line appears each in the control region (C), the IgG test region and IgM test region. The result is positive for IgG & IgM antibodies and may indicate an immune response to a COVID-19 infection.

**IgG POSITIVE:** Two lines appear. A colored line appears in the control region (C), and also in the IgG test region. The result may indicate the presence of the specific antibody produced by the immune system in response to a COVID-19 infection.

**IgM POSITIVE:** Two lines appear. A colored line appears in the control region (C), and also in the IgM test region. The result may indicate the presence of the general antibody produced by the immune system as a first response to a COVID-19 infection.

**NEGATIVE**: One colored line appears in the control region (C). No lines appear in the IgG and IgM test regions. This may indicate no immune response present for a COVID-19 infection

**INVALID**: Control line fails to appear. This could happen for multiple reasons. Please ensure that you still record and share the results so that the research team are aware.

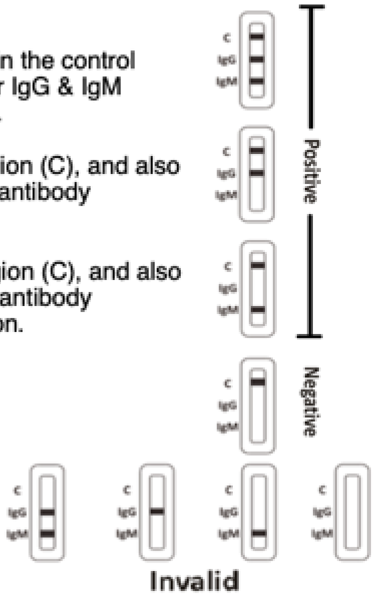


**Figure 1:** Result Interpretation

**Step 8: Record your result**

Without delay, place your test cassette in the box below and record your result, as well as the current time in ‘test end time’ (this should be in a 24-hour format, e.g. 17:05).

<table>
<tr>
<td>Place<br>test<br>cassette<br>here</td>
<td>Please complete the following based on your interpretation of the result:<br><br>Positive ☐<br>Negative ☐<br>Invalid ☐<br><br>Test start time: ____:____<br><br>Test end time: ____:____<br><br>«Login_ID» | 3 | 1</td>
</tr>
</table>

Take a photograph of the result. Make sure that the whole box is visible, and that the image is clear (see examples below). This must be done **within 10-15 minutes** of you completing the procedure to generate a valid result – **do not check or photograph the result after 20 minutes.**

✓ The whole box is clearly visible, with all information complete.

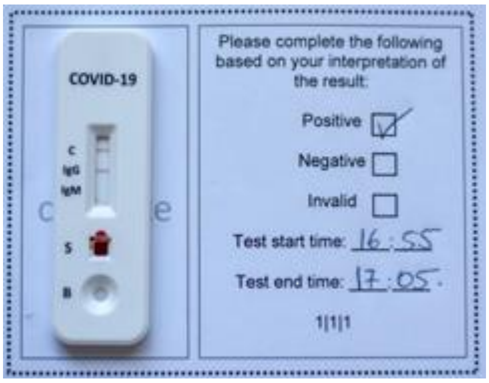


✗ Test obstructing information.

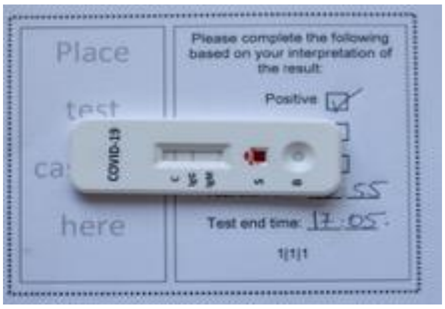


Whole box not visible.

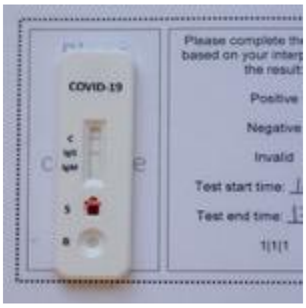


Image is blurry.

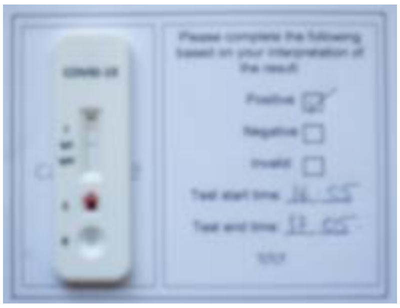

**Step 9: Upload your photograph**

Visit **www.kcl-check.org/test-results** via your web browser and upload and submit your photograph to share the test result with the KCL CHECK research team. You can use the submission form to let us know if you had any issue during the test. We would greatly appreciate if you could contact us via our study e-mail if you have been unable to upload an image of your result.

**Your Participant ID is: «Login_ID»**

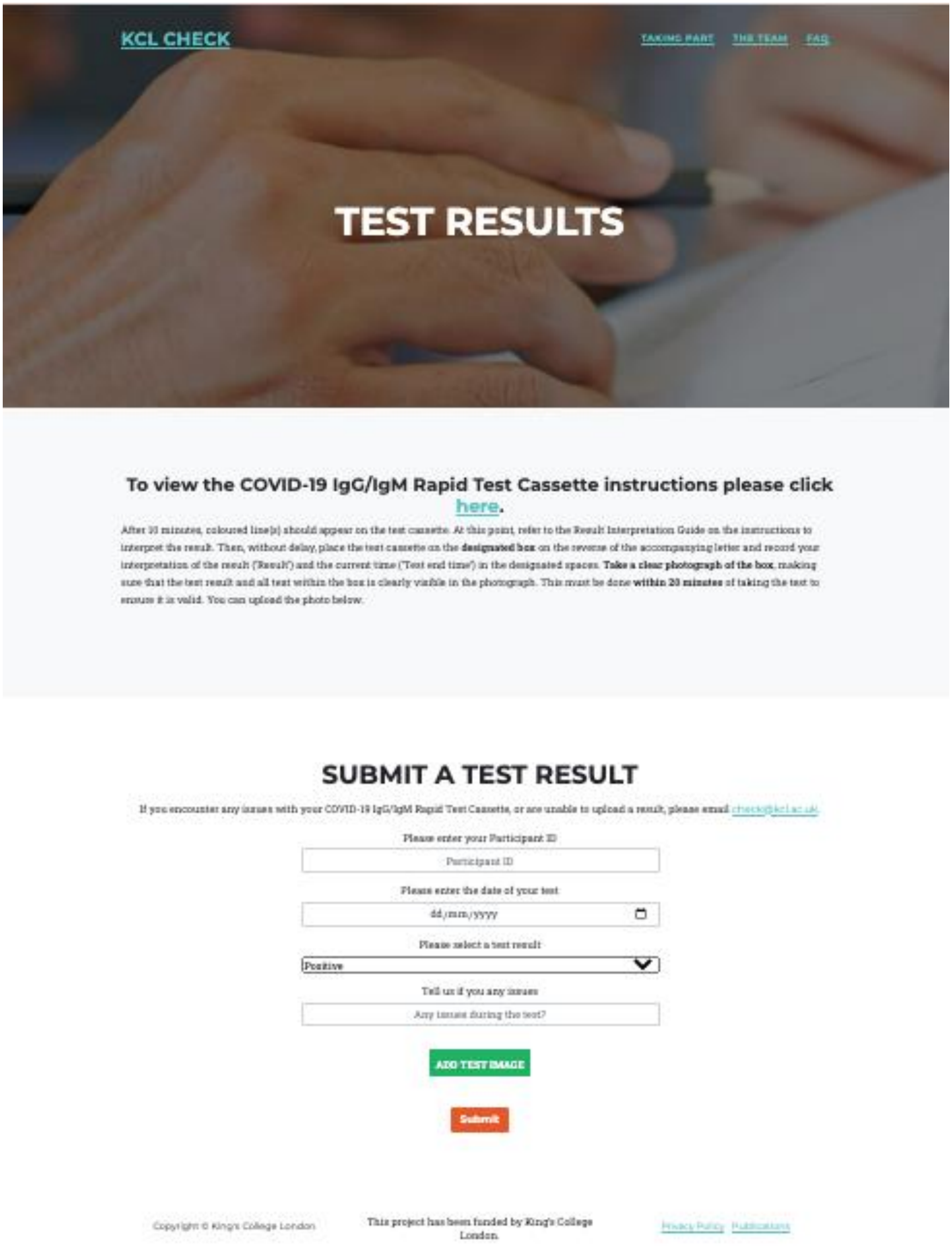

**Step 10: Dispose of the kit in the clear sample bag**

Place the used test cassette, buffer, lancet and alcohol wipe into the clear sample bag. **Seal the bag and dispose of in general waste.**
